# Using Micro-Cognition Biomarkers of Neurosystem Dysfunction to Define ADHD Subtypes: A Scalable Digital Path to Diagnosis Based on Brain Function

**DOI:** 10.1101/2023.01.22.23284871

**Authors:** Bruce E. Wexler, Ryan Kish

## Abstract

**Background:** Diagnostic categories in psychiatry are based on symptoms and include individuals with different underlying pathology. This within-diagnosis heterogeneity confounds new treatment development and treatment selection for individual patients. A research priority is to discover biomarkers that define groups of patients with similar neuropathology. Digital neurocognitive therapy (DNT) and assessment can provide micro-cognition biomarkers of unprecedented precision. The brain is hierarchically organized from single cells to neurosystems that integrate action of millions of neurons across the brain necessary for cognition and emotion. Micro-cognition biomarkers identify dysfunction at the level of neurosystems that produce clinical illness. We used micro-cognition biomarkers to identify subgroups of children diagnosed with ADHD but with different neuropathology.

**Methods:** K-means clustering was applied to 69 children 6-9 years old with ADHD using performance variables from a Go/NoGo test and the results analyzed against 58 typically developing (TD) children. Neurosystem dysfunction in each group was further characterized by micro-cognition biomarkers extracted from thousands of responses by each child during DNT.

**Findings:** Four highly reproducible clusters were identified that differed on emblematic features of ADHD. Cluster 4 showed two core ADHD features, poor response inhibition and inconsistent attention. Cluster 3 showed only poor response inhibition and the other two showed neither. Cluster 2 showed faster and more consistent responses, higher detection of simple targets and better working memory than TD children but showed the most marked performance decrements when required to track multiple targets or ignore distractors. Cluster 1 showed much greater ability recognizing members of abstract categories rather than natural categories that children learn through physical interaction with the environment while Cluster 4 was the opposite.

**Interpretation:** DNT provides data-rich fine-grained, low-cost, noninvasive, and scalable micro-cognition biomarkers that characterize subgroups of patients with the same symptom-based psychiatric diagnosis but differing neuropathology.

## Introduction

Diagnostic categories in psychiatry are currently based on clusters of symptoms and recognized to include individuals with different neuropathology. This heterogeneity arises from the fact that in different individuals, different neuropathology can lead to the same symptoms and the same neuropathology can cause different symptoms.^1–3^ These limitations in classification confound efforts to develop new treatments and select optimal treatments for individual patients. Accordingly, it is an important research priority to discover biomarkers of neuropathology that define homogeneous groups of patients with similar underlying neuropathology and characterize their pathology. We report that analysis of thousands of responses from individuals during digital neurotherapy can provide micro-cognition biomarkers of neurosystem dysfunction that identify clusters of children who share the diagnosis of ADHD but on the basis of different neuropathology.

ADHD has been called “an exemplar of a robust clinical neuropsychiatric syndrome with marked heterogeneity across multiple levels of analysis.”^4^ For example, multiple neuropsychological tests^4–6^, ratings of temperament and personality^7,8^, resting state connectivity on fMRI, clinical course^7^, and EEG power analyses^9^, all differentiate groups of children with ADHD from TD children, but each show abnormality in only a minority of children. Nearly 3,000 genetic studies, including 32 meta-analyses, demonstrate strong heritability but no defining features of ADHD per se.^10^ Using fMRI, clinical, and demographic data from multiple studies to differentiate ADHD from TD children, researchers identified a subgroup in which 94% of children had ADHD, but 80% of ADHD children were not in the group.^11^ Similar heterogeneity confounds diagnosis and treatment development in most brain disorders including depression, schizophrenia, autism, mild cognitive impairment, Parkinsonism and dementia.

In the present study we used objectively measured micro-cognition biomarkers to identify subgroups of children who share the diagnosis of ADHD but differ in neuropathology. The brain is hierarchically organized from single cells to cell dyads, local circuits of interconnected neighboring neurons and neurosystems that integrate the action of millions of neurons distributed across multiple brain regions.^12^ Biomarkers of function at the different levels are each of potential value and complement each other (Figure 1). Micro-cognition biomarkers assess function of neurosystems necessary for cognition and emotion. These are important since it is dysfunction at the level of neurosystems that alters thinking and feeling and produces clinical illness. Blood biomarkers provide information on pathology at cellular and synaptic function including inflammation and plasticity but are difficult to associate with brain physical or functional anatomy. Quantitative EEG and fMRI also provide information on neurosystems but are limited in specificity and scalability.

**Figure 1:**
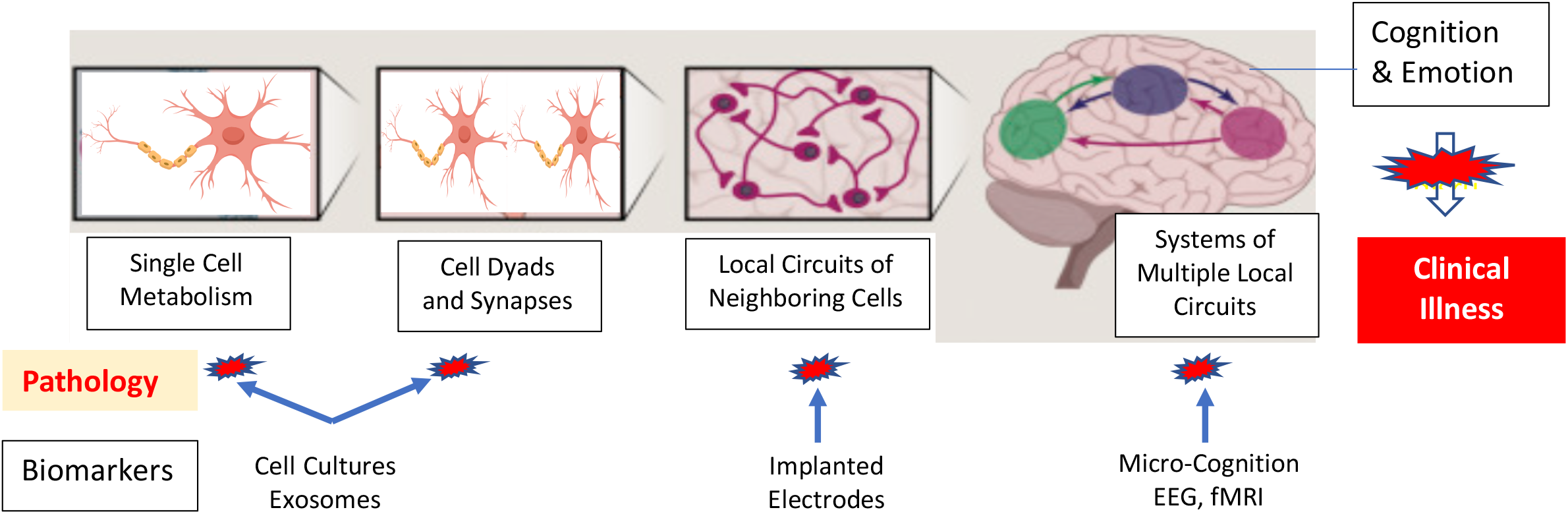
Brain hierarchical functional organization and biomarkers

Data for the present analyses derive from a study showing that digital neurotherapy (DNT) reduced symptoms and improved cognition in children with ADHD with the degree of symptom reduction associated with baseline and change measures of cognition.^13^ The DNT systematically challenges and trains incremental fine-grained variations in micro-cognitive elements of attention, response inhibition, speed of processing, working memory, use of categories, and pattern recognition across thousands of training trials, creating the opportunity to identify subgroups based on micro-cognition biomarkers. The DNT program also administers tests of response inhibition, focused attention, and working memory.^14,13^ We used measures of response inhibition and sustained attention from the Go/NoGo test of response inhibition, hallmark features of ADHD, in unstructured cluster analysis to identify potential subgroups of children. We then used the micro-cognition biomarkers generated by the DNT and scores on the focused attention and working memory tests to validate and further characterize neurosystem dysfunction in the clusters. TD children did the DNT program side-by-side with the ADHD children providing normative values.

## Experimental Methods

### Participants

Participants were recruited via letters to parents of children offering a free after school program to improve attention.^13^ All study procedures comply with ethical standards of the Human Investigation Committee at the Yale School of Medicine and with the Helsinki Declaration of 1975, as revised in 2008. Informed consent was obtained from parents or guardians, and all participants gave informed assent prior to study procedures. A child was considered “screen-positive” if the average rating per item was greater than 1·2 on the parent or teacher SNAP-IV rating scales^15,16^, and diagnosis confirmed using the Kiddie Schedule for Affective Disorders and Schizophrenia – Present and Lifetime Version administered by doctorate or master-level clinicians during interviews with parents. At the same time, the parent SNAP-IV was completed again facilitated by research staff. Diagnosis of ADHD was assigned if a child met DSM-IV TR criteria for ADHD or was deemed to be at “high-risk” for ADHD defined as one symptom below diagnostic criteria. Acute behavior problems, physical disability that would prevent them from participating in treatment or co-morbid psychiatric diagnosis except autism spectrum disorder were exclusion criteria. 91 children meeting criteria for ADHD and 86 children considered to be TD between 5–9 years old and IQ of at least 80 on the Kaufman Brief Intelligence Test (K-BIT^17^) were enrolled. Kindergarten children (n=18) were almost entirely TD younger siblings of other participants, enrolled for the convenience of parents and dropped from analysis to better match age in ADHD and TD children.

### Micro-Cognition Biomarkers Derived from Computer-Presented Cognitive-Training Exercises

The cognitive training games were designed by BEW and developed as web-based applications by the Yale startup company C8 Sciences. Children used three modules, each with 80–150 levels representing incremental changes in task features and difficulty that probe and train micro-variations in cognition and associated neurosystems. Every response from every child during training is captured for analysis. In the current analyses we used performance from the initial 150 minutes of training sessions only. The first module (CTB) begins with the child having to click on a yellow ball moving randomly across the screen whenever it turns red, exercising sustained attention. The ball moves faster following correct responses and slows after errors. After challenging simple target detection (level 1) by shorter durations of target color changes, the task changes so the ball sometimes turns blue (a foil, level 2) that is to be ignored, adding response inhibition. Next, the target color randomly changes back and forth between blue and red (level 3), increasing required response inhibition demands and adding cognitive flexibility. Two micro-cognition biomarkers were extracted: target detection rate (levels 1–3) and foil click rate (levels 2–3). In the second module (Fly), children click on butterflies carrying signs only if the object on the sign was a member of a designated category (letters, numbers, animals, plants, or food). With correct responses, the butterflies move faster and more butterflies appear at the same time. Performance measures are target detection rate in each category type at low or high processing loads (1–3 vs 4– 6 objects on screen). The third module (Ducks) requires the child to figure out the rule that links a series of three objects in linear array and use the rule to choose a fourth object from among three choices to complete the row. On some levels the patterns were sequences of colored shapes (e.g., red circle, yellow triangle, red circle, yellow triangle) and on others they were sequences of numbers (e.g., 1,1,2,2). Time to respond becomes shorter with correct responses. On some levels, the missing element in the pattern is in the last (fourth) position in the row (“what comes next” or wcn) and on others it is in positions 1, 2, or 3 in the row (“what comes before” or wcb). Extracted micro-cognition biomarkers were percent correct responses and average response times on correct responses.

### Formal Tests of Cognition

Three dimensions of executive cognitive function shown to be compromised in previous studies of children with ADHD—focused attention, response-inhibition, and working memory—were assessed with online versions of established research measures administered in the classroom setting. Classroom administration provides ecological validity of evaluation in the real-world learning environment but without the controlled environment of an office. As a result, we established validity criteria for each test based on accuracy on easier test trials and absence of too many responses that were either so fast or slow as to question whether the child was adequately engaged with and understood the task. Tests were given one per day beginning on the third day of the program. Two tests precisely followed the design from the NIH Toolbox of tests of executive function (nihtoolbox.org). The first was the Flanker Test of focused attention where the primary performance measures were percent correct and speed and consistency of reaction time on correct congruent and incongruent trials. In this task, children have to indicate by keyboard response the pointing direction (right or left) of the center arrow in a horizontal array of five arrows. On incongruent trials, the four “flanking” arrows point in the opposite direction of the central arrow. The test generated three primary performance measures: 1) Difference in percent correct between congruent and incongruent trials; 2) Difference in reaction time between correct congruent and incongruent trials; and 3) standard deviation of reaction time on correct congruent trials. The second test was the List Sorting Working Memory Test. Subjects are shown a series of animals or household objects and then click on the objects just seen in a grid of 16 objects in order from smallest to largest rather than the order in which they were presented. The test starts with a list of 2 objects. If the subject completes the list accurately, list length is increased by one. Two consecutive errors end the test. In part one, trials of animals and household objects alternate. In part two, animals and household objects are inter-mixed, and subjects have to reorder the animals first and then the household objects. Dependent measures were longest list lengths achieved in each part. The third test is a Go/NoGo test of response inhibition. Subjects are instructed to press the space bar whenever a “go” stimulus is presented but not when a “no-go” stimulus is presented. There are three blocks of 50 stimuli randomized in sets of 10 with 8 go and 2 no-go in each set. In the first block “P” is the go stimulus and “R” the no-go stimulus. In the second block this is reversed. In the third block, pictures of furniture are go trials and pictures of foods like cake and ice cream are no-go stimuli. Stimuli are presented for 400 msec with a 1400msec response window after stimulus offset. Errors are indicated by a large red “X.” Performance variables were: 1) percent of response to no-go trials; 2) response time on go trials; and 3) standard deviation of response times on go trials. Absences from the program on a day that a test was administered or failure to meet validity criteria led to variation in the number of children with data from each test.

### Statistical Analysis

Analyses used a limited set of variables with established relevance to ADHD in an initial unstructured K-means cluster analysis to identify clusters of ADHD children, evaluated cluster reproducibility in randomly generated split-half samples, and validated and further characterized clusters by comparison with the remaining test and micro-cognition variables. The three performance measures from the Go/NoGo test were used in the cluster analysis because they reflect two features often considered emblematic of ADHD: abilities to inhibit responses and to consistently maintain attention. 69 ADHD and 58 TD children had valid Go/NoGo tests and constituted the study sample for cluster and subsequent analyses (mean age (SD) for ADHD = 7·5 (1·1) and TD = 7·4 (1·1). At each stage of validation and further characterization, the clusters were compared to each other by one-way or mixed models repeated measures ANOVAs and relevant post-hoc tests. In addition, scores of the ADHD children were normed against those of TD children to determine whether specific strengths and weaknesses relative to other ADHD subtypes represented weakness or strengths compared to TD children. In addition, given the a priori overall study goal of identifying distinctive features of subgroups of patients sharing the clinical diagnosis of ADHD, and the robust reproducibility of the cluster categorizations (see results), significant differences between one cluster and the others suggestive of a defining feature of the cluster are noted even when the main effect of cluster considering differences between each cluster and all others did not reach significance. Only scores from valid tests were used when comparing clusters leading to variation in the number of subjects in different analyses.

## Results

### Cluster Definition and Replication

The three performance variables from the Go/NoGo test were normalized and entered into a K-means cluster algorithm with cluster number set to four by the “elbow” method. As seen in Figure 2, C4 shows two deficits often considered emblematic of ADHD, with response inhibition reflected in a high percent of responses on no-go trials and inconsistency of attention reflected in high within-subject standard deviation in response times on correct go trials, both > 2 SDs worse than TD children. C3 also shows a substantial failure of response inhibition but no increase in variability of response times. Strikingly, C2 children were faster and more consistent than TD in response time and consistency and essentially the same as TD in response inhibition. Finally, C1 children were marginally better than TD children in response inhibition but nearly 0·5 SD slower in response times suggesting good self-control and increased carefulness in response. These cluster differences were highly reproducible in two randomly generated split-half samples. The clusters did not differ significantly in age, but since C2 was older on average (8·1 +/− 1·2 years) than the others (C1, 7·5 +/− 0·8; C3, 7·3 +/− 1·1, and C4 7·1 +/− 1·3 years) differences between C2 and others reported below were confirmed in analyses with age as a covariate.

**Figure 2:**
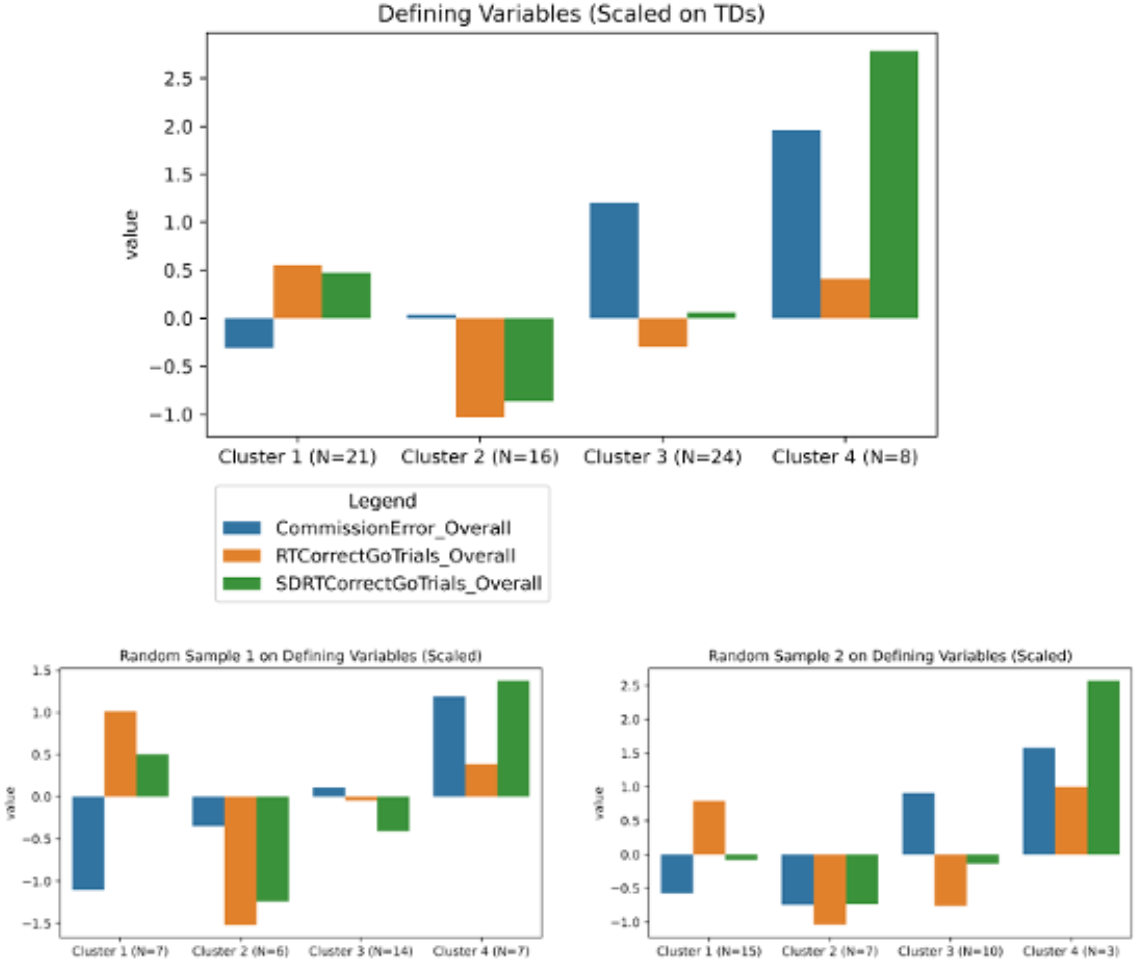
Four clusters of children with ADHD defined on the basis of failure to inhibit responses to no-go trials (commission errors), response time on go trials, and standard deviation of response times on go trials.

### Comparison of the Clusters on the Flanker and LSWM Tests

Flanker Test: None of the one-way ANOVAs for the three performance variables showed significant differences across clusters. However, inspection of the data showed that the impact of the flanking/distracting arrows on response time (response time correct incongruent – response time correct congruent) was nearly one-half SD greater (0·46) in C2 than in TD children while it was actually smaller in C1 and C4 than in TD and nearly the same as TD in C3 (figure 3A). The t-test between C2 and the other clusters combined was significant (t[63]=2·03, p=0·047) while the differences among C1, C3 and C4 did not approach significance.

**Figure 3:**
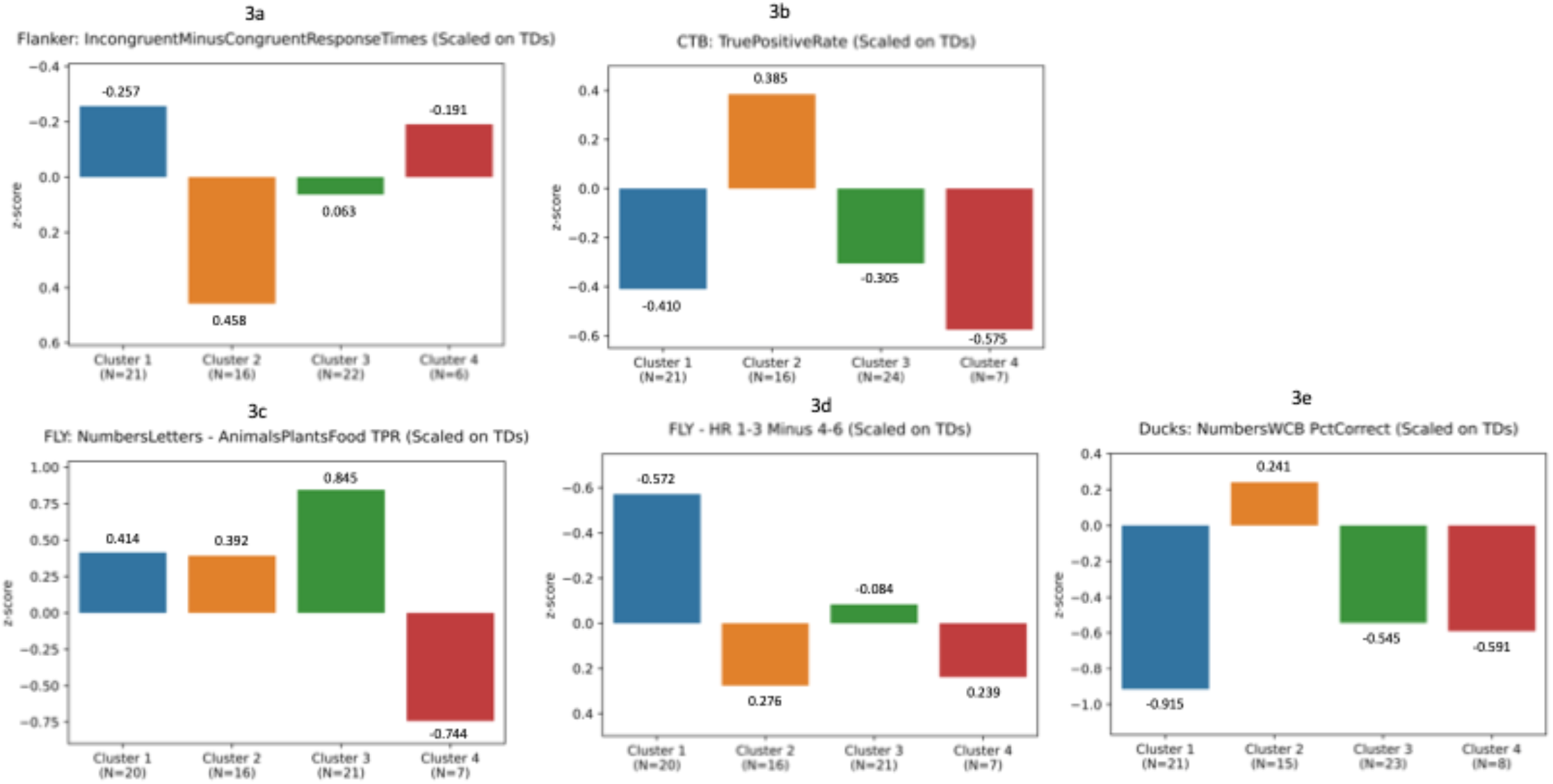
Comparisons of clusters on micro-cognrtion performance variables. Values are standard deviations from typically developing children. Values below the zero line represent performance worse than typical developing except in the 2c where the vanable is the difference in performance when assigning objects to abstract categories versus natural categories. All four clusters showed greater differential performance than typically developing children, but the difference was particularly pronounced in C3 and C4 but with opposite directionality in the two clusters.

**Figure 4:**
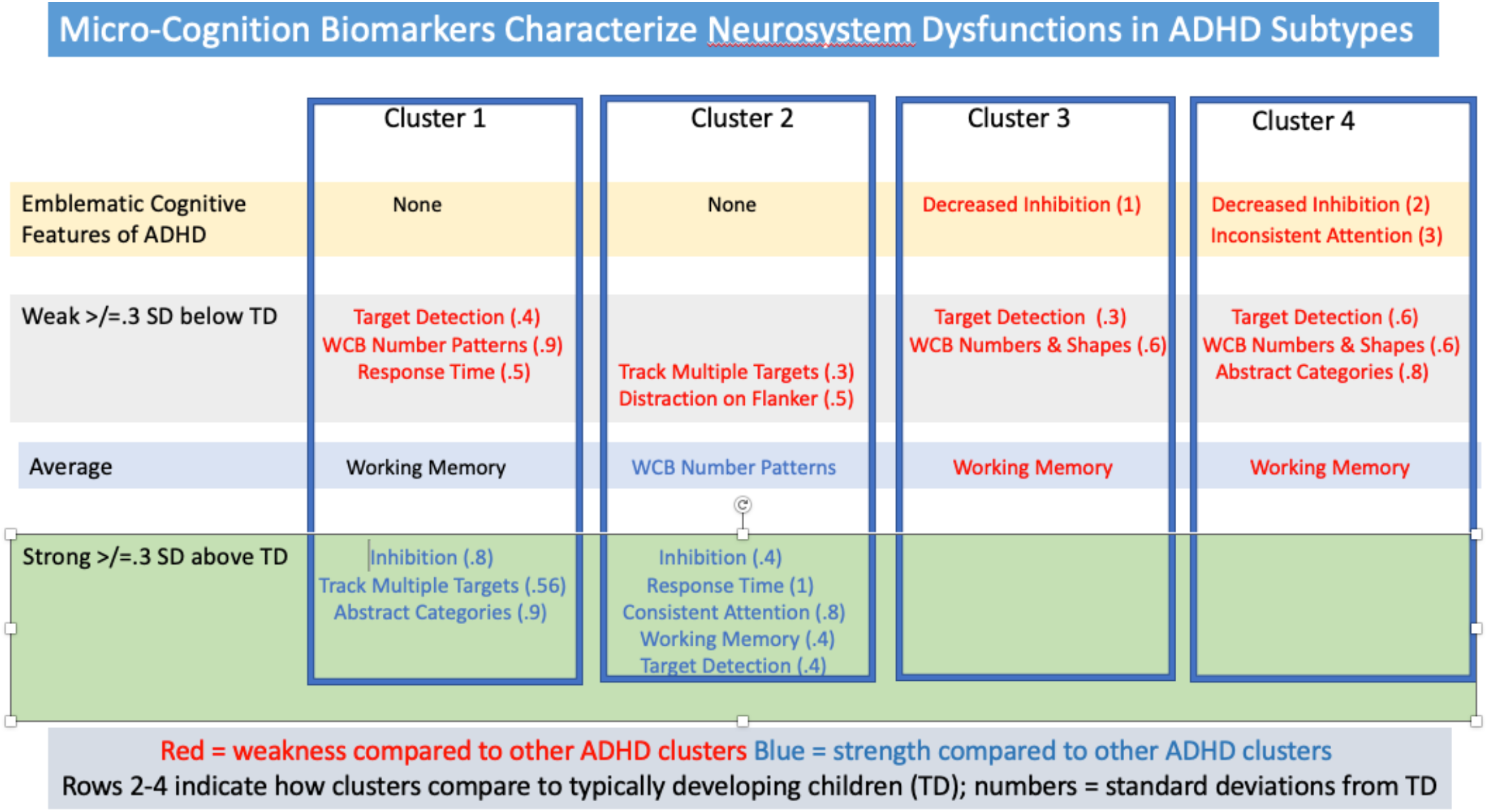
Overview of differences in micro-cognition biomarkers of neurosystem dysfunction among four subtypes of ADHD. Inhibition, Consistent Attention, and Response Time come from Go/NoGo Test, Distraction from Flanker Test, Working Memory from LSWMT, Target Detection from CTB sustained attention DNT module, Multiple Target Cost and Abstract vs. Natural Categories from Fly category DNTmodule, What Comes Before (wcb) pattern recognition from Ducks pattern recognition DNT module.

LSWM Test: A mixed model ANOVA with cluster as the between subjects factor and longest list length achieved in levels one and two as within-subject repeated measures showed main effects of cluster (F[3, 50]=2·85, p=0·046) and level (F[1, 50]=19·37, p<0·0001) while the interaction did not approach significance. Independent t-tests on the average of maximum list length on the two parts showed the main effect of cluster was due to significantly better performance by C2 compared to C3 and 4 (t[35]=3·31, p=0·0022).

### Micro-Cognition Biomarkers Derived from DNT Cognitive-Training Exercises

#### CTB Module

One child from C4 was excluded from analysis due to responding in the absence of a target or foil stimulus 10 times as frequently as the average of all others. A mixed model ANOVA with cluster as the between subject factor and level (simple target detection, target with foils, and switching target) as a within-subject factor yielded significant main effects of cluster (F[3, 64]=3·07, p=0·034) and level (F[2, 128]=6·15, p=0·0028), with C2 performing better than all others (C2 vs. C1: t[35]=3·17, p=0·0031; C2 vs. C3: t[38]=2·46, p=0·018; C2 vs. C4: t[21]=2·39, p=0·026) and better than TD (figure 3B), and level 3 being more difficult than the first two levels. Similar analysis of foil click rate showed significantly higher rates on level 3 (t[134]=5·75, p<0·0001) but no significant differences among clusters or interaction.

#### Fly Module

A mixed model ANOVA with cluster as the between subject factor and category (numbers, letters, animals, plants, and food) and difficulty (either 1–3 or 4–6 items on the screen) as within-subject factors yielded significant main effects of cluster (F[3, 60]=3·67, p=0·017), category (F[3, 194]=15·05, p<0·0001), and difficulty (F[1, 60]=36·44, p<0·0001), a significant interaction of cluster x category (F[9·71, 194·24]=4·48, p<0·00083), and a trend level interaction of cluster x difficulty (F[3, 60]=2·26, p=0·090).

The main effect of cluster reflected lower overall performance by C4 (accuracy 93% +/− 4·9%) compared to the other three which did not differ significantly (C1 97·6% +/− 1·89%, C2 96·7% +/− 3·0%, C3 96·5% +/− 3·5%). The robust cluster x category interaction was driven by relative differences between performance on numbers and letters—abstract human made categories—compared to performance on plants, animals, and food which are natural categories that children interact with physically.^18^ Combining numbers and letters, and plants, animals, and food, the interaction between cluster and category type was significant as expected (F[3, 60]=7·99, p<0·00014). C3’s preferential ability on numbers and letters is 0·85 SD greater than TD while C4 differed from TD by 0·75 SD in the opposite direction due to higher performance identifying animals, plants, and food (figure 3C).

Given the large differences between C4 and the others in overall performance, and our *a priori* interest in evaluating the differences among the other clusters in micro-cognition, we repeated the mixed model ANOVA with only clusters 1–3. The cluster effect was now non-significant (F[2, 54]=0·72, p=0·49) while the cluster x difficulty interaction was significant (F[2, 54]=3·14, p=0·05); C2 showed the largest decline in performance from low to high difficulty on all five categories and C1 showed the smallest decline on all five categories with C3 in the middle (figure 3D). This distribution of frequency in a 3×3 table of clusters and relative rank in performance decrement is significant by Chi-Square (chi-square=30, df=4, yates corrected p=0·0007).

#### Ducks Module

A mixed model ANOVA of correct responses with cluster as the between subject factor and category (colors/shapes or numbers) and position of missing element (wcb or wcn) as within-subject factors yielded a non-significant main effect of cluster (F[3, 63]=1·92, p=0·14), significant main effects of position (F[1, 63]=26·87, p<0·0001) and category (F[1,63]=8·57, p=0·0048), and significant interactions of cluster x position (F[3, 63]=3·40, p=0·023) and cluster x position x category (F[3, 63]=2·95, p=0·040). The three-way interactions was driven by higher performance in C2 than the other three clusters (p-values ranged from 0·0012 to 0·0088) on the numbers wcb level (figure 3E).

A similar ANOVA using response time on correct responses as the performance variable yielded significant main effects of cluster (F[3, 63]=3·62, p=0·018), position (F[1, 63]=4·46, p=0·039) and category (F[1,63]=7·46, p=0·0081) without significant interactions. C4 was slower in response times compared to the other clusters (C4 vs C1 +2 +3 t[65]=2·564, p=0·013), while across all clusters responses were slower on wcb than wcn trials and to colors/shapes patterns than to number patterns.

## Discussion

Diagnosis in psychiatry and many other brain disorders is based on clusters of symptoms that appear by consensus and tradition to have similar form, whereas as diagnosis and treatment in other branches of medicine advanced when knowledge of the normal physiology of the organ system involved provided a logic to diagnose and objective measures to assess disease^1^. Identification of biomarkers of brain dysfunction holds promise to provide a similar path forward for brain disorders. Pathology can exist at any of the hierarchical levels of brain functional organization, and biomarkers at each level are needed. We demonstrate the value of digital micro-cognition biomarkers of the neurosystems pathology that leads to alterations in cognition and emotion that constitute clinical illness. Micro-cognition biomarkers are generated by a DNT program that collects thousands of responses from each patient while probing and training fine-grained incremental variations in perceptual and cognitive processing. The small incremental variations in neurosystem processing demands provide high precision and sensitivity. The micro-cognition biomarkers are low cost, non-invasive and scalable.

Model-free cluster analysis identified four clusters of children all sharing the diagnosis of ADHD but differing in hallmark features of ADHD—response inhibition and speed and consistency of response. The clusters were highly consistent in randomly generated split-half samples even in the relatively small overall sample. Comparison of clusters on the List Sort Working Memory Test, the Flanker Test, and multiple micro-cognition biomarkers extracted from responses to incremental variation in DNT training trials provided external validation of the clusters and initial steps in characterizing neurosystem pathology in each.

Figure 3 summarizes the distinguishing micro-cognition strengths or weaknesses in each cluster relative to the other clusters and TD children. C1 children were slower in response than C2–C4 and TD, and had lower target detection on fast moving targets and decreased ability to recognize patterns of numbers displayed in left-to-right sequences than C2 and TD when the missing element was in one of the first three positions in the sequence (“what comes before, wcb”) rather than the last position (“what comes next, wcn”). On the other hand, C1 had stronger self-control, showed less performance decrement when having to categorize 4–6 objects on the screen compared to 1–3 objects, and showed a much greater performance advantage when identifying members of the abstract categories of numbers and letters compared to the natural categories of animals, plants and food compared both to other ADHD clusters and TD. Together these features suggest a slower and more controlled cognitive style, keeping their efforts, speed, and physicality within a comfort range. Perhaps their ADHD-like symptoms are related to slow response times giving the impression of distraction.

C2 children had two areas of deficit compared both to TD and at least two other ADHD clusters. These otherwise cognitively strong children were particularly impacted when challenged by having 4–6 objects moving on the screen compared to 1–3 objects during categorization training and when distracted by arrows pointing in the opposite direction of the central target arrow on the Flanker Task. These weaknesses are striking in contrast to their consistency of attention and superior performance on simple target detection tasks (Go/NoGo and CTB training module) compared both to other ADHD and TD children. Both can be understood as limits in ability to screen out distractions not present in the simpler target detection tasks. In the Flanker Test, the four distracting arrows are presented 30msec before the target adding primacy to the distraction. In the category training module, distraction is both the concurrent internal categorization task and associated changes in neurosystem configuration as the type of category changes, and the external perceptual load. Together these findings suggest highly efficient and focused attention in many situations, but an abrupt drop off when the full field of simultaneous perceptual and cognitive processing exceeds a capacity point. Inability to attend sufficiently when the threshold is exceeded produces symptoms meeting current criteria for diagnosis of ADHD, but the dysfunction is very different from those in the other clusters despite the shared symptom-based diagnosis. These children may be the ones recognized as dually exceptional—abilities and limitations—or high functioning ADHD.

C3 children showed decreased inhibitory control which is probably the basis of their ADHD diagnosis but without evidence of inconsistent attention, the other hallmark symptom of ADHD. While other differences between C3 and other clusters and TD were limited, they were more challenged by the “what comes before” condition compared to C1, C2, and TD on the pattern recognition training task with both numeric and geometric shape patterns. The wcb condition requires holding an “empty” space in mind and circling back to fill it in. This requires the neurosystems to configure for both pattern recognition and more robust working memory and attention demands, as well as what is involved in the empty space representation. Their weakness on both types of patterns rather than only in numeric patterns differentiates them from C1, suggests dysfunction in aspects of neurosystem configuration challenged by the wcb task configuration in general, rather than C1’s more fine-grained deficit limited to wcb recognition of sequences of numbers. It is possible that the task-general requirements of inhibitory control, maintenance of an “empty space” representation, and increased working memory demand combine at the neurosystem level to produce the deficit.

C4’s performance was more than two SDs worse than TD children in both response inhibition and ability to maintain attention consistently on the Go/NoGo test. In addition, when required to identify objects belonging to specific categories, they had more difficulty picking out letters or numbers than animals, plants, or foods while the other three clusters showed the reverse pattern. Animals, plants, and food are categories developed on the basis of real-life sensory experience while numbers are abstract human-made categories based more on frontal executive cognitive function. Perhaps these children have a more physical interaction with their environments and may therefore need types of learning opportunities not offered in traditional education settings.

Identification of subgroups of children whose ADHD symptoms are associated with different underlying neurosystem dysfunctions is important for matching patients with the treatment most likely to benefit them and guiding new treatment development. Children with some neurosystems dysfunctions, for example, may benefit from one type of medicine, DNT, behavioral or psychotherapy but not another. While the micro-cognition biomarkers used in the present report provide only a limited characterization of the pathology in each cluster, they demonstrate the feasibility and potential power of the approach. As the work developing and applying micro-cognition biomarkers proceeds, more complete information about the nature of neurosystem pathology in different clusters of patients from different current symptom-based diagnostic categories will enable more complete depictions of pathology. As with other research about the nature of CNS pathology, the information will also provide new insights into normal brain functional organization based on dysfunctions that co-occur, and potentially provide new concepts that better integrate the types of biomarker abnormalities seen in each of ADHD clusters. Comparison with serum biomarkers, fMRI, and EEG of groups of patients who, on the basis of shared neurosystem dysfunction as revealed by micro-cognition biomarkers are more pathologically homogeneous, will further characterize the nature of different brain disorders and aspects of normal brain organization. Reverse translation of the neurosystem dysfunctions characterizing patient subgroups will provide clinically relevant animal models for rational drug development.

The study has several limitations. First the overall study sample is not very large, and one cluster included fewer than 10 subjects. In addition to general instability inherent in small samples, power to demonstrate statistical significance in comparison of the smallest cluster to the others is limited and may lead to underestimation of differences. The different cognitive tests and micro-cognition biomarkers might vary in sensitivity and reliability leading to appearance of deficit specificity when instead it is a manifestation of a less sensitive marker failing to show a significant difference while a more sensitive one does. While study data included biomarkers of multiple aspects of cognition and neurosystem function, there are many additional aspects of cognition not assessed but needed for fuller characterization of the cluster-specific pathology. Measures related to spatial working memory are notable among additional potentially valuable measures as it has been shown to be compromised in ADHD. To realize the potential suggested by the present study, replication in larger samples and longitudinal use in studies comparing different treatments are needed.

## Conclusion

Micro-cognition biomarkers of neurosystem dysfunction and unstructured data analytic methods identified four groups of children all of whom shared the same clinical diagnosis but differ in neurosystem dysfunctions. The findings provide proof-of-concept for use of micro-cognition biomarkers to refine current diagnostic procedures. These improvements are needed to better match patients with treatments and define pathophysiologically homogeneous patient samples for developing new treatments and characterizing neuropathology. With regard to the ADHD subtypes, additional studies and larger study samples are needed to replicate and identify additional features of each cluster before an integrated view of the system dysfunction can be developed. Comparison of the clusters with serum biomarkers of neuropathology and functional brain imaging activation paradigms informed by the identified differences in micro-cognition are needed to build models of different pathologies. Reverse translation to create clinically relevant animal models can potentially increase the percentage of candidate drugs that prove effective in clinical trials, and lead to rationally developed combinations of pharmaco- and behavioral therapies. Finally, while our DNT will require additional testing for validation, a digitally-delivered objective methodology for better classifying ADHD and other brain disorders offers the possibility of a remote assessment, treatment, and longitudinal tracking solution that can increase access to care and allow clinicians to treat more patients more effectively at lower cost.

## Data Availability

All data upon which the analyses reported in the present study are available upon reasonable request to the corresponding author.

## Contributions

BEW conceived of the project, designed the digital neurotherapy program and platform that acquired the data, helped design and interpret the data analysis, and was the primary writer of the manuscript. RK collected the data, cleaned the data, prepared the data, helped design the data analysis, produced the statistical analyses, produced the figures, and interpreted results. Both authors were involved in drafting and revising of the document.

